# Epidemiological profile, trend, seasonality, and factors associated with Yellow fever disease and barriers to vaccination: a retrospective observational study of multi-season national surveillance data in Nigeria, 2017 - 2021

**DOI:** 10.1101/2023.09.20.23295820

**Authors:** Stephen Eghelakpo Akar, William Nwachukwu, Sunbo Oludare Adewuyi, Anthony Agbakizua Ahumibe, Iniobong Akanimo, Oyeladun Okunromade, Chinwe Ochu, Olajumoke Babatunde, Ifedayo Adetifa, Chikwe Ihekweazu, Mami Hitachi, Kentaro Kato, Yuki Takamatsu, Kenji Hirayama, Satoshi Kaneko

## Abstract

Since its resurgence in 2017, Yellow fever (YF) outbreaks have continued to occur in Nigeria despite routine immunization and implementation of several reactive mass vaccination campaigns, resulting in substantial morbidity and mortality. Nigeria is considered a high-priority country for implementing the WHO EYE strategy, which is targeted at eliminating YF outbreaks by 2026. This retrospective observational study was conducted to describe the epidemiological profile of reported cases, trends, and seasonality of YF incidence; identify factors associated with Yellow fever disease (YFD) and barriers to YF vaccination in Nigeria. Univariate, bivariate and multivariate binary logistic regression analysis was done. Of 13014 suspected YF cases, 7640 (58.7%) had laboratory confirmation for Yellow fever virus (YFV). Predictors of YFD were male sex (aOR 2.36, 95% CI: 1.45-3.91) compared to female; age group being 15-29 years (aOR 4.13, 95% CI: 1.59-13.00) compared to under-five; residing in the Derived Savannah (aOR 30.10, 95% CI: 11.50-104.00), Lowland/Mangrove/Freshwater rainforest (aOR 8.84, 95% CI: 3.24-31.10), Guinea Savannah/Jos Plateau (aOR 6.13, 95% CI: 1.90-23.50) compared to the Sahel/Sudan savannah; working in outdoor settings compared to indoor (aOR 1.76, 95% CI: 0.96-3.22); and vomiting (aOR 2.62, 95% CI: 1.39-4.83). The rainy season was protective against YFD (aOR 0.32, 95% CI: 0.19-0.52) compared to the dry season. Because being unvaccinated emerged as protective factor (aOR:0.51, 95% CI: 0.25-1.00) compared to those with unknown vaccination status, the data was further disaggregated by vaccination status. Predictors with higher odds ratios were found among unvaccinated. Predictors of YFD among the vaccinated were the first quarter compared to the second quarter of the year (aOR 4.04, 95% CI: 1.48-12.95) and residing in the southern region compared to the north (aOR 14.03, 95% CI: 4.09-88.27). Barriers to YF vaccination were the rainy season compared to the dry season (aOR 1.29, 95% CI: 1.05-1.57), being 15 years or older (15-29: aOR 2.06, 95% CI: 1.51-2.83; 30-44: aOR 2.11, 95% CI: 1.45-3.07; 45-59: aOR 2.72, 95% CI: 1.63-4.58; 60+: aOR 6.55, 95% CI: 2.76-17.50), residing in the northern region (aOR 3.71, 95% CI: 3.01-4.58) compared to the south, and occupation being butcher/hunter/farmer (aOR 2.30, 95% CI: 1.52-3.50) compared to home-based/office workers. Being a student was protective against being unvaccinated (aOR 0.62, 95% CI: 0.47-0.83). Several factors were associated with YFD, which were aggravated by lack of vaccination. Although barriers to vaccination were elucidated, inadequate vaccination coverage alone may not account for the recurrent outbreaks of YF in Nigeria. These findings are critical for planning public health interventions and to guide further research that would enable Nigeria end YF epidemics.

## Introduction

Yellow fever (YF), an arthropod-borne, re-emerging viral hemorrhagic illness, caused by the Yellow fever virus (YFV), has remained a significant public health concern in Africa and the Americas, accounting for about 29,000 to 60,000 deaths annually [1–8]. An effective and safe vaccine against YFV, the live attenuated YF 17D vaccine, has been available for over seventy years and integrated into routine immunization (RI) program in endemic countries. The vaccine has been shown to confer 95% protection in vaccinated populations within 30 days of vaccine uptake and offers life-long immunity [5,6,9–13]. Infected persons present with a broad spectrum of clinical disease, ranging from asymptomatic infections and mild febrile illness to severe disease and death. It is estimated that 15-25% of infected symptomatic persons progress to severe disease with a case fatality rate of 20-60% [6,14].

Despite the existence of RI for YF and the implementation of several mass preventative and reactive vaccination campaigns, YF outbreaks have continued to occur in Nigeria since its resurgence in 2017. Previous studies in Nigeria have focused on the descriptive epidemiology of outbreak cases [6,11] and vaccination program implementation [5,15]. Tobin *et al.* (2021) assessed the knowledge and attitudes towards YF in a cross-sectional study conducted in a small community in one of the hotspots states in southern Nigeria [16]. There is paucity of studies on the factors associated with YFD and YF vaccination in Nigeria. The trend and seasonality of YF in Nigeria has not also been adequately described.

The Eliminate Yellow fever Epidemics (EYE) strategy of the World Health Organization is targeted at eliminating YF outbreaks in endemic countries by the year 2026, and Nigeria is considered a high priority country for its implementation [17]. The EYE strategy is aimed at protecting at-risk population, preventing international spread of YF, and containing local outbreaks of YF promptly [17]. The objective of this study is to describe the epidemiological profile of reported YF cases, trends, and seasonality of YF incidence; as well as identify factors associated with YFD and barriers to YF vaccination in Nigeria. This has become imperative as public health authorities aim to develop strategic public health interventions to enable the country to meet the 2026 target of the WHO EYE strategy.

## Methods

### Data Source Description

We retrospectively analyzed YF surveillance data obtained from the Surveillance, Outbreak, Response, Management, and Analysis System (SORMAS) platform managed by the Nigeria Centre for Disease Control (NCDC) to which State ministries and Local Government departments of health reported. SORMAS in Nigeria, part of the integrated disease surveillance and response system (IDSR), has been previously described [5,11]. In brief, once a patient meets the standard case definition for a suspected case of YF, the attending physician notifies the Local Government Area (LGA) Disease Surveillance and Notification Officer (DSNO); the LGA DSNO then notifies the State Epidemiologist and the State DSNO, who verifies the case and then inform the focal person at the NCDC. The NCDC, in conjunction with the State and the LGA officers, constitutes an outbreak investigation team to investigate the case using a case investigation form (CIF). The cases are line-listed, and blood samples are collected for laboratory confirmation in line with the National Guidelines for YF Preparedness and Response [5,18]. Serological testing involved the detection of YF-specific antibodies in the serum samples of suspected cases using Enzyme-linked Immunosorbent Assay (ELISA; U.S. CDC MAC protocol; done in-country) and plaque reduction neutralization test (PRNT; done at the Regional Reference Laboratory, Dakar, Senegal). ELISA positive results were recorded as “presumptive positive” while negative results were recorded as “negative cases”. Both ELISA positive samples and “inconclusive samples” were sent to the Regional Reference Laboratory for confirmation by PRNT. Molecular testing by real-time polymerase chain reaction (RT-PCR) was done in-country for samples collected from suspected cases within ten days of onset of symptoms to detect YF viral genome in blood. PCR negative samples were further tested by serology to rule out false negative results while positive samples are reported as confirmed YF cases [18]. Information on the CIF is logged into the SORMAS platform using the epidemiological number issued by the State Epidemiologist as a unique identifier. The data abstracted from the SORMAS platform include epidemiological number, socio-demographic information (age, sex, education, occupation), vaccination status, clinical information (date of onset, date of admission, symptoms), location (state), trave history, and laboratory test status (negative, positive, unconfirmed) from July 2017 to December 2021.

### Case definition

The case definitions applied for YF surveillance in Nigeria are as follows: A suspected case of YF was defined as “any person with acute onset of fever or history of fever and/or general body weakness, vomiting, bleeding from any part of the body, and jaundice appearing within 14 days of onset of the first symptom” [18]. A presumptive case was defined as a “suspected case in whom was found the presence of YF IgM antibody in the absence of YF vaccination within the last 30 days before onset of illness or positive post-mortem liver histopathology or epidemiological link to a confirmed case or an outbreak”. A confirmed case was defined as “ presumptive case with one of the following: detection of YF IgM; detection of four-fold rise in IgM or IgG antibody titers between acute and convalescent serum samples or both; detection of YF-specific neutralizing antibodies and absence of YF immunization within 30 days before onset of illness or one of the following: detection of YF viral genome in blood, liver or other organs by immunoassay, isolation of YF virus and absence of YF immunization within 14 days before onset of illness [18]. However, for this study, a positive case was defined as a suspected case that was confirmed positive for YFV by either serology or RT-PCR. A negative case is a suspected case that was confirmed negative by both serology and RT-PCR. An unconfirmed case is a suspected case for which laboratory test result was missing in the line list.

### Additional data sources for the analysis

Administratively, Nigeria has 774 LGAs, which are clustered into 36 states and the Federal Capital Territory and zoned into six geo-political zones (North-Central, North-East, North-West, South-East, South-West, and South-South). The geo-political zones were created based on political and socio-cultural considerations from the northern and southern regions. There are two climatic seasons in Nigeria (dry season and rainy season). The rainy season begins in April and ends in October while the dry season commences from November and terminates at about the end of March [19]. Forests spanned over nine ecological zones [20] (**S1 Fig**). The northern part of the country is dotted with Savannah-type vegetation whereas the southern region is characterized by weather conditions that promote luxuriant rainforest and proliferation of insect populations. It is estimated that forest reserves occupy 10% of the total land mass, with 78.7% in the Savannah zone, 7.7% in the derived Savannah zone, and 13.6% in the rain forest zone [19,21]. The ecological zones have been described as running from the northern to the southern parts in a contiguous fashion, in which natural ranges extending from the mangrove swamps and tropical forests in the coastal flanks become shrubs and Savannah woodlands in the low Plateau, which extends into the semi-arid plains of the northern region in central Nigeria and the eastern highlands. This complex mesh of forest pattern had resulted in a variety of vegetation in the landscape, including Sahel, Guinea, and derived Savannah as well as lowland rain forest, freshwater and mangrove forests, coastal marshes, and estuaries. The ecological zones are repositories of a wide variety of plants and animal species, including 20,000 insect and 247 mammalian species [19]. The variables geo-political zone, geographical region, and ecological zone were generated based on the states in which cases were reported.

### Definition of variables

Time-oriented variables such as annual quarter, epidemic period (pre-pandemic, pandemic) and climatic season (dry season, rainy season) were generated by grouping cases by date of onset of illness. Epidemic period was defined as whether a case developed symptoms before (pre-pandemic) or during (pandemic) the period of the COVID-19 pandemic (January 2020 and beyond) while climatic season was defined as whether the case occurred during the months of the dry season or the rainy season. Socio-demographic factors presumed to be associated with outcome variables were identified from previous studies [2,4,22]. Age in years and sex were analyzed as reported by cases and relatives. Age was used to create two sets of age group variables: For the analysis of factors associated with outcome variables (YFD and being unvaccinated) without disaggregation, age was categorized into seven levels (0-4, 5-9, 10-14, 15-29, 30-44, 45-59, and 60+ years); however, when YFD was disaggregated by vaccination status, age was categorized into two levels (<15 and >=15 years) to accommodate small sample sizes. Geo-political zone, geographical region, and ecological zone were generated using the state in which cases were detected. Educational status was created from the educational level reported by cases – primary, secondary, tertiary, Islamic education, no education. These were collapsed into two levels: all those who had any formal education were defined as “educated” and those with no education were defined as “no education” alongside those who did not report their educational status, considering the current illiteracy level of nearly 40.0% in Nigeria. Occupational group was generated from the occupation reported by cases, which were loosely defined. Those working in office settings, housewives, and kids below school age were grouped as “home-based/office workers”; those who reported business, trading, plumbing, electrician etc. were grouped as “artisans/traders”. In Nigeria the term “business” loosely includes people involved in any form of buying and selling of goods and services. Cases who were of school age (6-17 years) but were not reported as students were grouped as “students” alongside those who reported being students based on the premise that basic education is mandatory by law in Nigeria. Those involved in animal sales, butchers, hunters, and farmers were grouped as “butchers/farmers/hunters”; and those working in healthcare settings were defined as health workers. Cases who reported no occupation were defined as unemployed in view of the high unemployment rate in Nigeria.

### Statistical analysis

De-identified and anonymized national line-list of YF outbreak cases from 1st July 2017 to 31^st^ December 2021 was abstracted from the SORMAS platform in compliance with NCDC data sharing protocol. We received the dataset in Microsoft Excel format from the data management on 5^th^ February 2022 with personal identifiers of the cases removed. Categorical variables were summarized using frequencies and percentages while numerical variables were summarized using median and inter-quartile range. Our main outcome variables were: (1) YFD, defined as a suspected case of YF confirmed for YFV by either serology or RT-PCR (a categorical variable with two levels: No = No YFD, Yes = YFD) and (2) unvaccinated, defined as a suspected with no previous YF vaccination before the onset of symptoms (a categorical variable with two levels: No/Yes). Comparison of percentages between outcome and explanatory variables was done using Pearson’s Chi square statistic and Fisher’s exact test for categorical variables and Kruskal-Wallis rank sum test for numerical variables with p-value <0.05 considered to be statistically significant. We used date of onset of illness to plot epidemic curves with trend lines (histogram with line plots) and time series seasonality plots (line plot in which cases were aggregated by months over the entire study period) to show trends of cases over time. A population-adjusted age-sex pyramid of cases was also plotted using the ages of cases and country age-sex population structure.

At bivariate analysis, odds ratio (OR) was used as a measure of association between seasonal, geographical, socio-demographic, and clinical characteristics and respective outcome variables (YFD: No = 0, Yes = 1; unvaccinated: No = 0; Yes = 1) and the 95% confidence intervals with p-values (<0.05) were estimated. Subsequently, multivariate logistic regression analysis was done using stepwise selection process, which was based on Akaike Information Criterion (AIC) [23] - implemented in R using the *stepAIC* function in the MASS package [24] with backward selection. Here, explanatory variables with level of significance p<0.15 at bivariate analysis were included in the multivariate logistic regression model to determine adjusted odds ratios, 95% confidence interval, and p-values of predictors of the respective outcomes. We used variance inflation factor to assess multicollinearity of the variables in the final model and variables that exhibited collinearity values >= 5.0 were excluded from the final model. For variables that exhibited collinearity, the one with a higher variance inflation factor was dropped from the final model. For both outcome variables, bivariate and multivariate analysis was first done with the whole dataset and then disaggregated by vaccination status (YFD) and laboratory test result (being unvaccinated). We conducted our analysis with RStudio version 4.4.2 and R version 4.4.2 (http://posit.co/download/rstudio-desktop/). The main R packages used in our analysis were the tidyverse package (data wrangling) [25], tableone package (descriptive table) [26], gtsummary and finalfit packages (bivariate and multivariate analysis)[27], and TSstudio package (seasonality analysis)[28].

### Ethical consideration

Approval for this study was sought by submission of the study proposal (see Other supporting material) to the Research Ethics Committee of the Nigeria Centre for Disease Control and Prevention, which was reviewed and approved. The Research Ethics Committee then requested the authors to complete, sign, and submit a data sharing agreement form to access the dataset (see Other supporting material). The data has been previously collected, stored, and analyzed as part of outbreak response activity by the NCDC during which a case investigation form was administered by trained clinical staff to obtain relevant information from suspected cases of YF to which they consented in writing following detailed explanation about the use of their information by appending their signature and witnessed by an attending nurse and a family relative. In the case of minors, informed consent was given by either a parent or guardian. The data was confidentially stored in passworded computer systems by NCDC data management team and was shared in de-identified and anonymized Microsoft Excel format for this study. This necessitated a waiver of the need for informed consent for this study by the Research Ethics Committee.

## Results

### Trend and seasonality of case incidence

Case reporting was higher during the 2019 epidemic season (34.0%) while YFV positive case was higher during the 2020 epidemic season (44.0%). While suspected case reporting was higher during the third and fourth quarter of the year, YFV positivity was more in the fourth quarter of the year (**Table 1**).

**Table 1.**
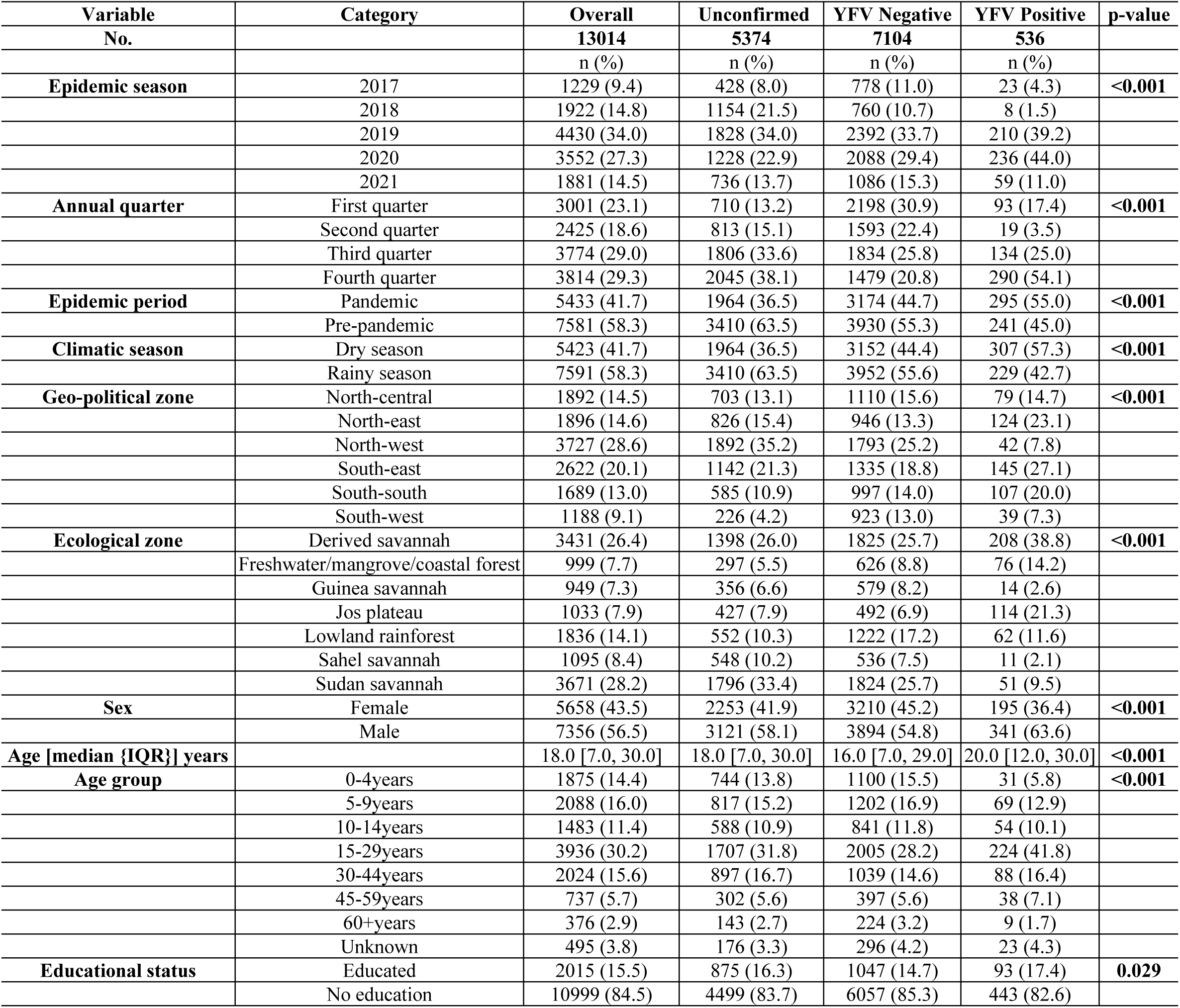
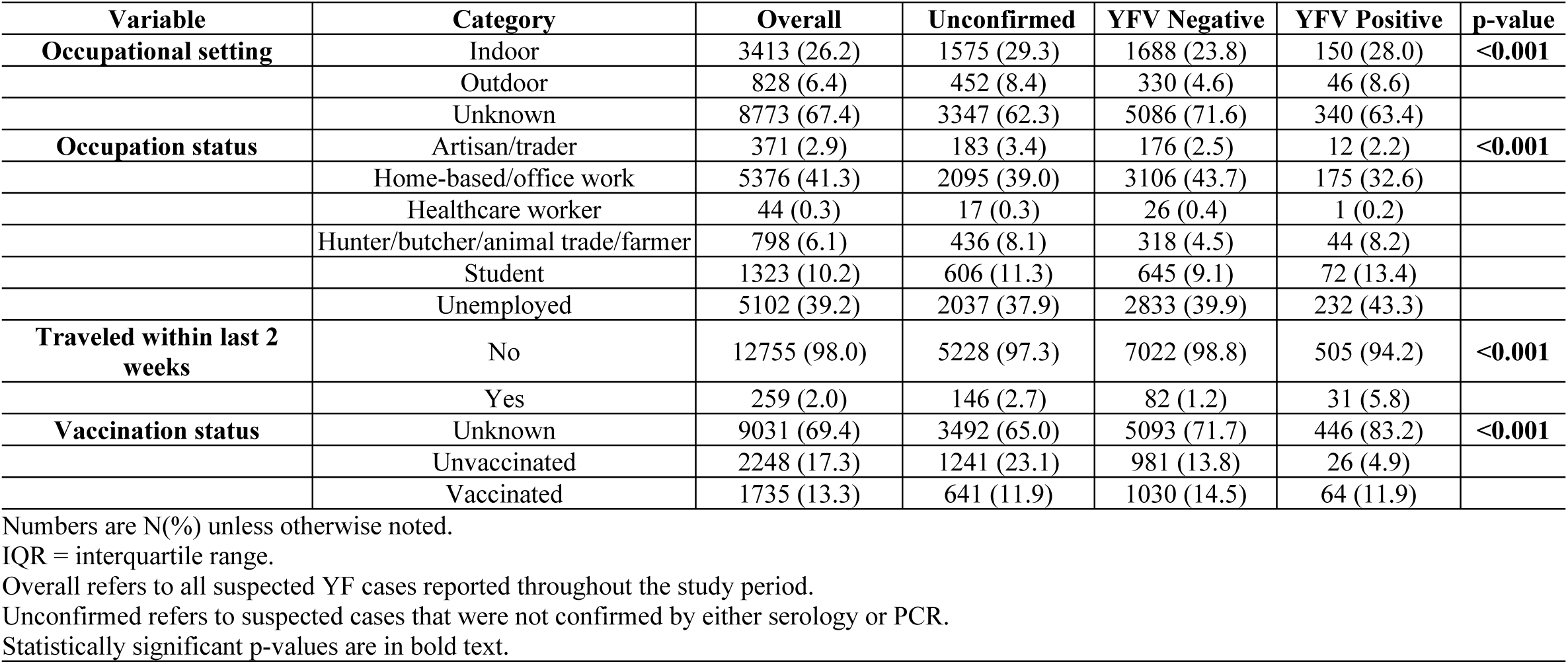
Seasonal, geographical, and socio-demographic characteristics of YF cases in Nigeria, 2017 and 2021 compared by laboratory test status.

The highest peak of case reporting and YFV case positivity were seen around Week 43 in the 2020 epidemic season (**Fig 1A**). The highest incidence of involvement of vaccinated cases was seen during week 34-43 of the 2019 epidemic season (**Fig 1B**). The initiation of YF vaccination (reactive and preventive) campaign between 2017 and 2018 apparently resulted in a flattening of the 2018 epidemic peak (around week 35-40).

**Fig 1.**
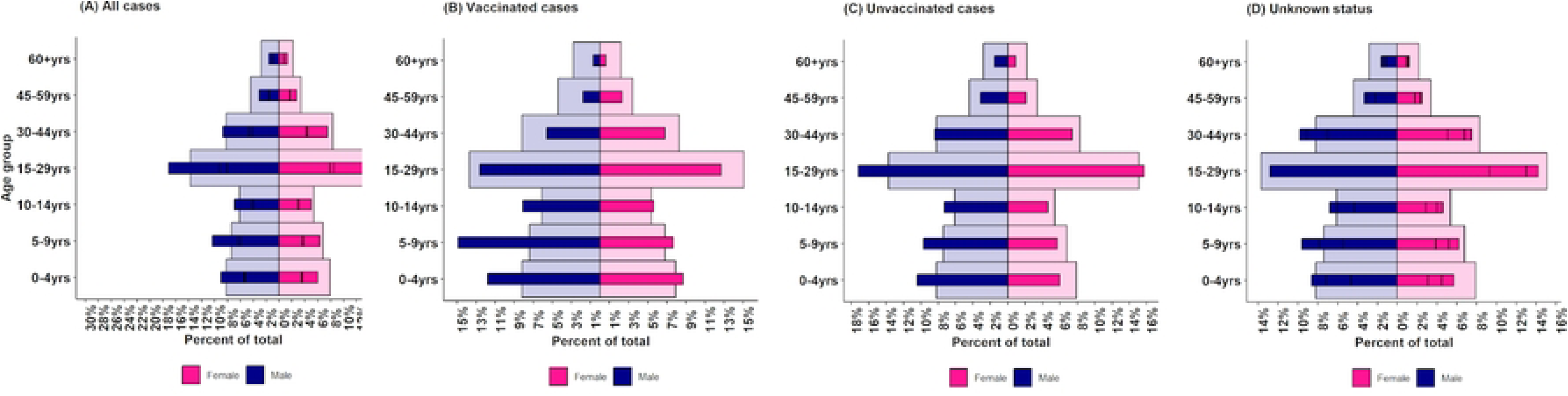
Time trends of incidence of YF cases in Nigeria, 2017-2021. Lines indicate weekly mean incidence. In: (A) average weekly incidence were stratified by laboratory test result; (B) average weekly incidence disaggregated by vaccination status. Light blue histogram background is the epidemic curve of all suspected cases.

The North-west geo-political zone presented a somewhat characteristic trend of case incidence pattern with several peaks seen over the entire period, usually during the dry season (highest peak seen between week 34 and 43 of the 2019 epidemic season), which were much larger in magnitude and longer in duration, except during the 2020 epidemic season (Fig 2A). This pattern was found to be inherent in the Sahel Savannah ecological zone (Fig 2B).

**Fig 2.**
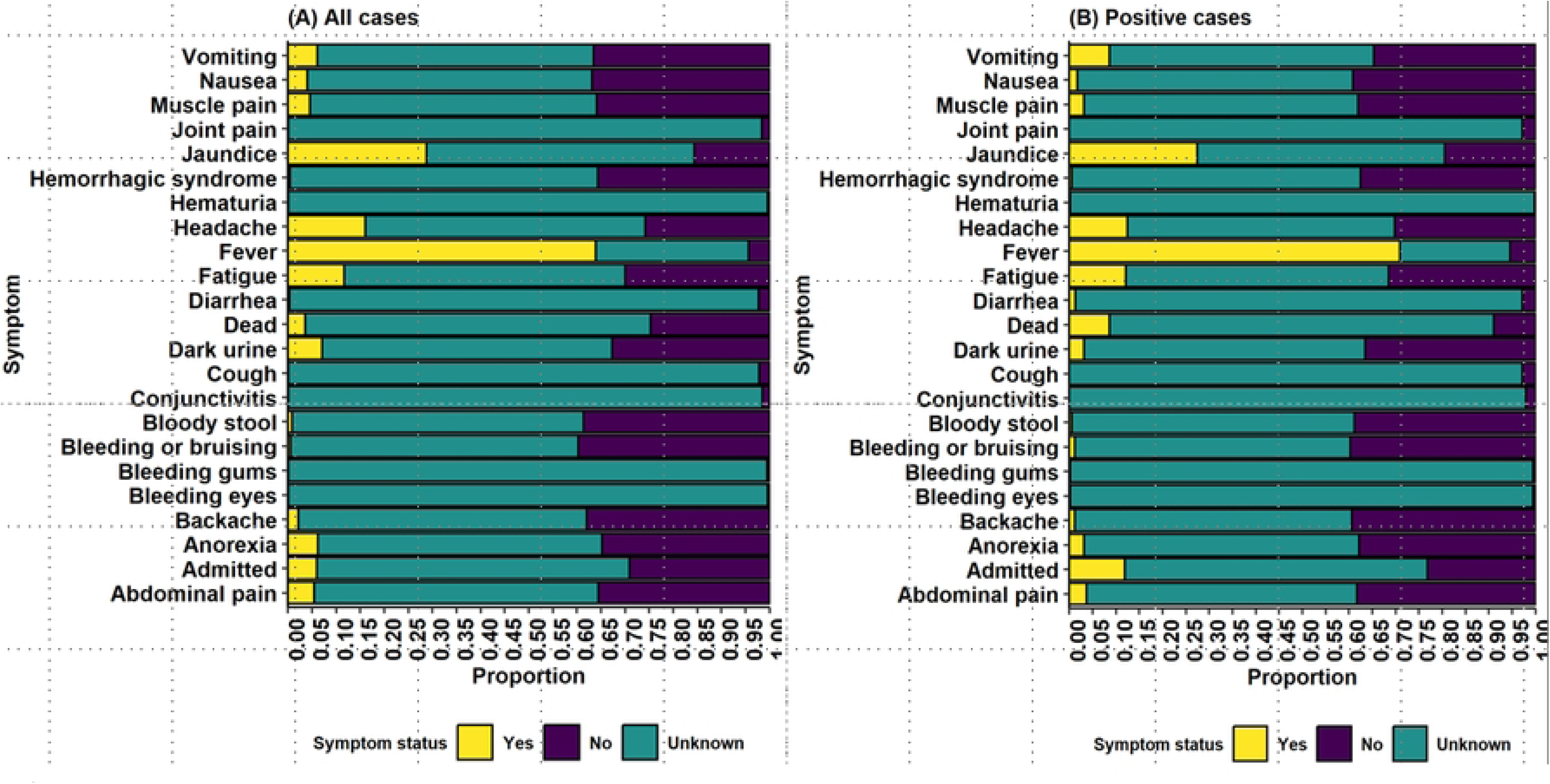
Trends of incidence of reported YF cases in Nigeria, 2017-2021. Lines indicate weekly mean incidence. In: (A) cases were disaggregated by geo-political zone; and (B) cases were disaggregated by ecological zone. Light blue histogram background is the epidemic curve of all reported cases.

The 2020 epidemic season presented the most distinct trend pattern throughout the entire period in which the highest peak incidence seen in the South-east geo-political zone reflected the outbreak pattern of the Derived Savannah. Annually, YF case reporting increased progressively from about the middle of the rainy season between June and July with the peak incidence seen sometime between September and November (Figs 3A and B).

**Fig 3.**
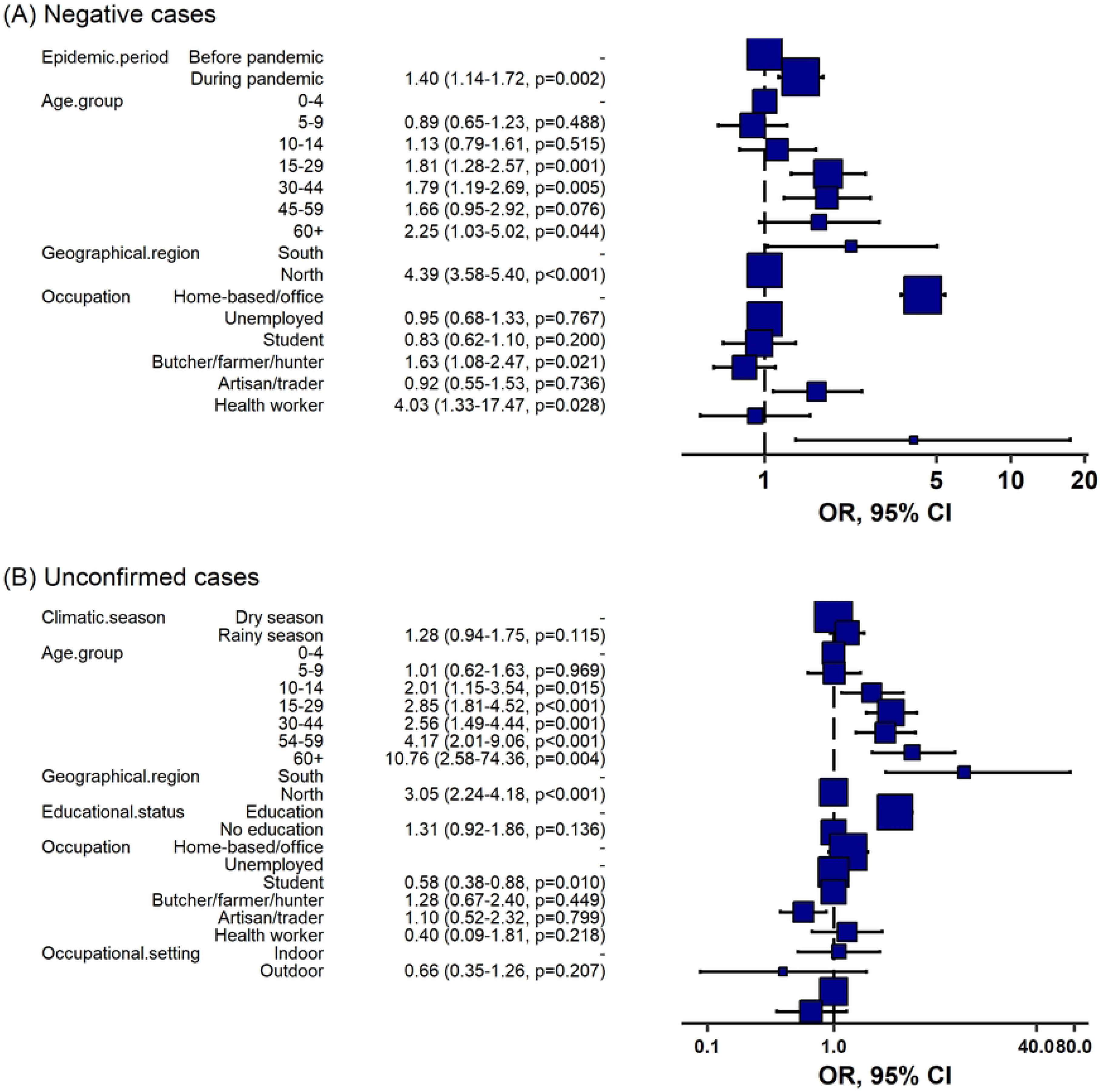
**Seasonality plot of reported Yellow fever cases in Nigeria, 2017-2021**. (A) all suspected cases, (B) YFV confirmed cases. Cases were aggregated by month in a line plot over the entire period.

Overall case reporting was higher during the pre-pandemic era (2016-2019; 58.3%) while YFV positive case was more during the COVID-19 pandemic era (2020-2021; 55.0%; <0.001) (Table 1).

### Socio-demographic and clinical characteristics

Between July 1, 2017 and December 31, 2021, Nigeria recorded a total of 13,014 suspected YF cases, of which 7640 (58.7%) had laboratory confirmation by either serology or RT-PCR. Overall YFV case positivity was 7.02% (536/7640). Median age was higher among YFV positive cases (median= 20.0 years; IQR= 12.0 – 30.0 years) (Table 1). The most affected age group were those 15-29 years, who also accounted for about 30.0% (64,883,373) of the total population and about 32.0% (3936) of all suspected cases (Fig 4A). They were also the most unvaccinated group (Fig 4C) and had the largest proportion of persons with unknown vaccination status (Fig 4D).

**Fig 4.**
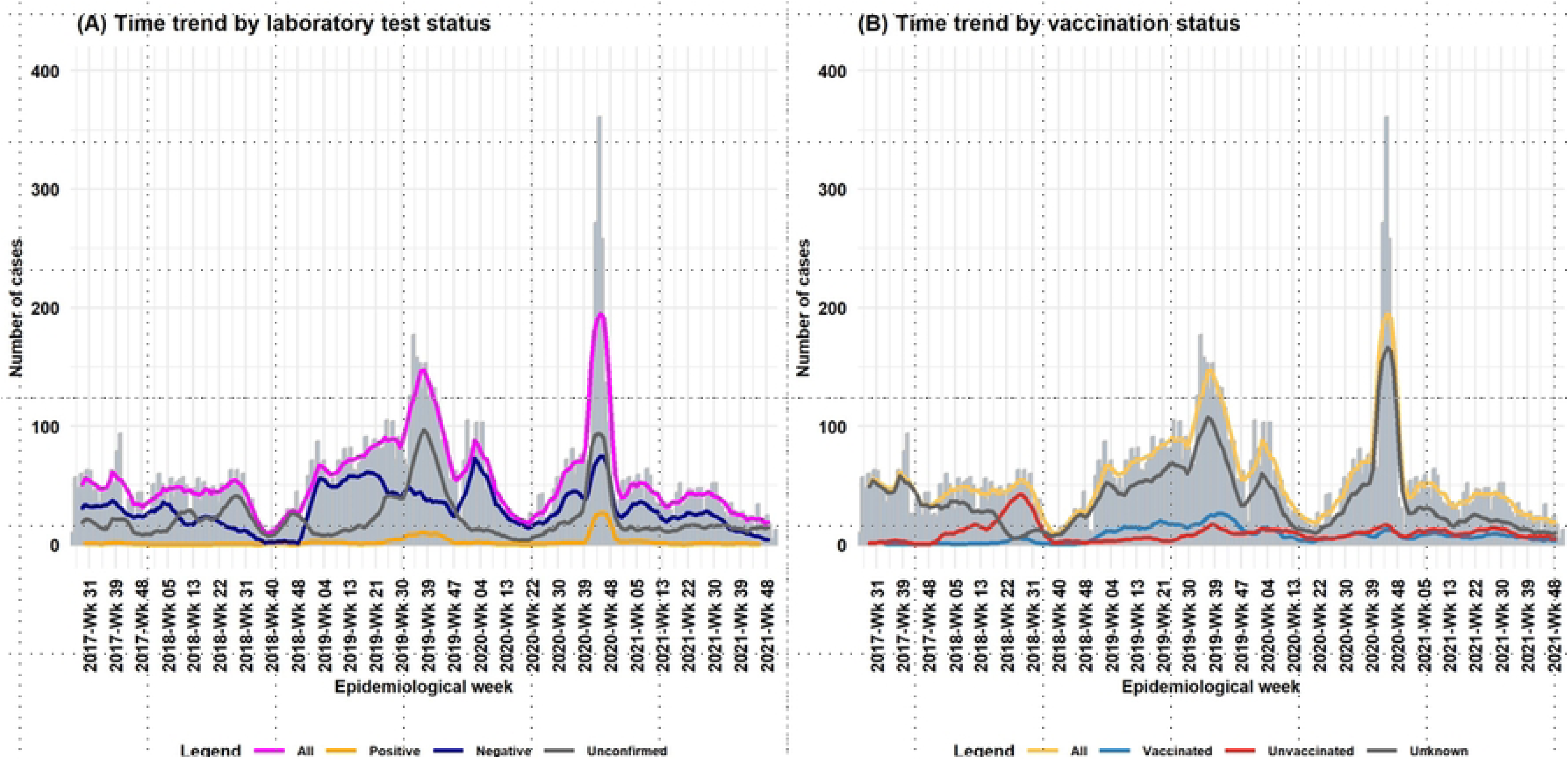
Age-sex pyramid of Yellow fever cases in Nigeria, 2017-2021. From left to right: (A) all reported cases, (B) vaccinated cases, (C) unvaccinated cases, and (D) cases of unknown vaccination status. The percentage of cases were superimposed on country age-sex population structure. Pink = female and blue = male. The light-shaded bars indicate country demographic age-sex structure.

Cases were mostly males (all: 56.5%; positive: 63.6%; negative: 54.8%; unconfirmed: 58.1%; p<0.001). Case reporting was higher in the Sudan Savannah (28.2%) whereas YFV positive case was more in the Derived Savannah (38.8%, p<0.001) ecological zone. They were predominantly of uneducated (all: 84.5%; positive: 82.6%; negative: 85.3%; unconfirmed: 83.7%; p=0.029); mostly home-based or working in office setting (41.3%), but the unemployed had higher YFV case positivity (43.3%, p<0.001). They were largely of unknown vaccination status (all: 69.4%; positive: 83.2%; negative: 71.7%; unconfirmed: 65.0%; p<0.001) (Table 1).

Clinical symptoms commonly presented among reported cases were fever (63.9%), jaundice (28.7%), headache (16.1%), fatigue (11.6%), dark urine (7.1%), and vomiting (6.1%). YFV positive cases presented more with fever (70.9%) and had a higher admission rate (11.9%) (Fig 4).

**Fig 5.**
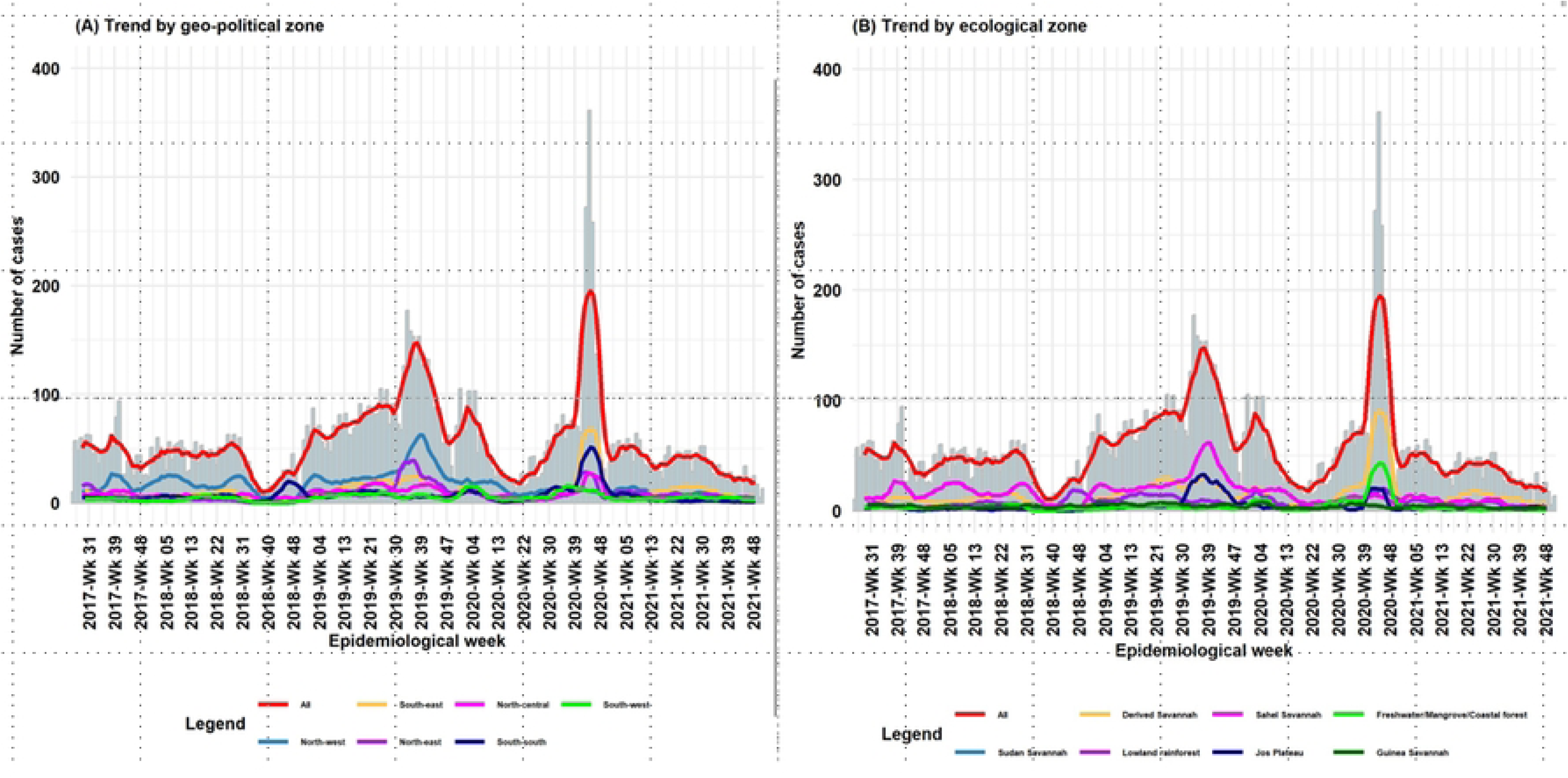
Clinical characteristics of reported Yellow fever cases in Nigeria, 2017-2021. (A) all reported cases, and (B) YFV confirmed cases. Yellow bar = symptom is present (Yes), purple bar = symptom not presented, and green bar = missing data.

### Seasonal, geographical, socio-demographic, and clinical factors associated with YFD

Factors associated with YFD are shown in Table 2. Independent predictors of YFD were male sex compared to female (aOR: 2.36, 95% CI: 1.45–3.91; p<0.001); age group being 15-29 years compared to under-fives (aOR: 4.13, 95% CI: 1.59–13.00, p=0.007); residing within the Derived Savannah, Lowland/Mangrove/Freshwater rainforest, and the Guinea Savannah/Jos Plateau compared to the Sahel/Sudan Savannah (aOR: 30.10, 95% CI: 11.50–104.00; aOR: 8.84, 95% CI: 3.24–31.10; aOR: 6.13, 95% CI: 1.90–23.50 respectively, p<0.001). Working in outdoor setting was associated with YFD but not statistically significant (aOR: 1.76, 95% CI: 0.96–3.22, p=0.067). Clinical predictor of YFD was presenting with vomiting (aOR: 2.62, 95% CI: 1.39–4.83; p=0.002). The rainy season was protective against YFD (aOR: 0.32, 95% CI: 0.19–0.52, p<0.001).

**Table 2.** Seasonal, geographical, socio-demographic, and clinical factors associated with YFD among reported cases in Nigeria, 2017-2021.

|  | Univariate |  |  |  | Multivariable |  |  |
| --- | --- | --- | --- | --- | --- | --- | --- |
|  | N=7640 |  |  |  |  |  |  |
| Characteristic | Total | Crude OR | 95% CI | p-value | Adjusted OR | 95% CI | p-value |
| **Epidemic season** | 7,640 |  |  |  |  |  |  |
| 2017 |  | 1.00 | Ref | <0.001 |  |  |  |
| 2018 |  | 0.36 | 0.15, 0.77 |  |  |  |  |
| 2019 |  | 2.97 | 1.96, 4.72 |  |  |  |  |
| 2020 |  | 3.82 | 2.53, 6.07 |  |  |  |  |
| 2021 |  | 1.84 | 1.14, 3.06 |  |  |  |  |
| **Annual quarter** | 7,640 |  |  |  |  |  |  |
| First quarter |  | 3.55 | 2.21, 6.01 | <0.001 |  |  |  |
| Second quarter |  | 1.00 | Ref |  |  |  |  |
| Third quarter |  | 6.13 | 3.87, 10.3 |  |  |  |  |
| Fourth quarter |  | 16.4 | 10.6, 27.2 |  |  |  |  |
| **Epidemic period** | 7,640 |  |  |  |  |  |  |
| Pre-pandemic |  | 1.00 | Ref | <0.001 |  |  |  |
| Pandemic |  | 1.52 | 1.27, 1.81 |  |  |  |  |
| **Climatic season** | 7,640 |  |  |  |  |  |  |
| Dry season |  | 1.00 | Ref | <0.001 | 1.00 | Ref | <0.001 |
| Rainy season |  | 0.59 | 0.50, 0.71 |  | 0.32 | 0.19, 0.52 |  |
| **Geo-political zone** | 7,640 |  |  |  |  |  |  |
| North-west |  | 1.00 | Ref | <0.001 |  |  |  |
| North-central |  | 3.04 | 2.09, 4.49 |  |  |  |  |
| North-east |  | 5.60 | 3.94, 8.10 |  |  |  |  |
| South-east |  | 4.64 | 3.30, 6.66 |  |  |  |  |
| South-south |  | 4.58 | 3.21, 6.67 |  |  |  |  |
| South-west |  | 1.80 | 1.16, 2.81 |  |  |  |  |
| **Ecological zone** | 7,640 |  |  |  |  |  |  |
| Sahel/Sudan Savannah |  | 1.00 | Ref | <0.001 | 1.00 | Ref | <0.001 |
| Derived Savannah |  | 4.34 | 3.27, 5.84 |  | 30.10 | 11.50, 104.00 |  |
| Lowland/Freshwater/Mangrove rainforest |  | 2.84 | 2.10, 3.88 |  | 8.84 | 3.24, 31.10 |  |
| Guinea Savannah/Jos Plateau |  | 4.55 | 3.35, 6.25 |  | 6.13 | 1.90, 23.50 |  |
| **Sex** | 7,611 |  |  |  |  |  |  |
| Female |  | 1.00 | Ref | <0.001 | 1.00 | Ref | <0.001 |
| Male |  | 1.42 | 1.19, 1.71 |  | 2.36 | 1.45, 3.91 |  |
| **Age group** | 7,321 |  |  |  |  |  |  |
| 0-4 years |  | 1.00 | Ref | <0.001 | 1.00 | Ref | 0.007 |
| 5-9 years |  | 2.04 | 1.34, 3.18 |  | 1.94 | 0.65, 6.57 |  |
| 10-14 years |  | 2.28 | 1.46, 3.61 |  | 1.96 | 0.60, 6.99 |  |
| 15-29 years |  | 3.96 | 2.75, 5.92 |  | 4.13 | 1.59, 13.0 |  |
| 30-44 years |  | 3.01 | 2.00, 4.63 |  | 2.36 | 0.76, 8.39 |  |
| 45-59 years |  | 3.40 | 2.09, 5.57 |  | 2.84 | 0.79, 11.2 |  |
| 60+ years |  | 1.43 | 0.63, 2.92 |  | 1.33 | 0.15, 8.38 |  |
| **Educational status** | 7,640 |  |  |  |  |  |  |
| Educated |  | Ref | 1.00 | 0.100 |  |  |  |
| No education |  | 0.82 | 0.66, 1.04 |  |  |  |  |
| **Occupational group** | 7,640 |  |  |  |  |  |  |
| Home/office worker |  | 1.00 | Ref | <0.001 |  |  |  |
| Unemployed |  | 1.45 | 1.19, 1.78 |  |  |  |  |
| Student |  | 1.98 | 1.48, 2.63 |  |  |  |  |
| Butcher/hunter/farmer |  | 2.46 | 1.71, 3.45 |  |  |  |  |
| Artisan/trader |  | 1.21 | 0.63, 2.12 |  |  |  |  |
| Healthcare worker |  | 0.68 | 0.04, 3.24 |  |  |  |  |
| **Occupation setting** | 2,214 |  |  |  |  |  |  |
| Indoor |  | 1.00 | Ref | <0.001 | 1.00 | Ref | 0.067 |
| Outdoor |  | 2.30 | 1.71, 3.11 |  | 1.76 | 0.96, 3.22 |  |
| **Traveled within last 2 weeks** | 7,640 |  |  |  |  |  |  |
| No |  | 1.00 | Ref | <0.001 |  |  |  |
| Yes |  | 5.26 | 3.40, 7.94 |  |  |  |  |
| **Vaccination status** | 7,640 |  |  |  |  |  |  |

Table 2 Cont.
| Characteristic | Univariate |  |  |  | Multivariable |  |  |
| --- | --- | --- | --- | --- | --- | --- | --- |
|  | Total | Crude OR | 95% CI | p-value | Adjusted OR | 95% CI | p-value |
| Unknown |  | 1.00 | Ref | **<0.001** | 1.00 | Ref | 0.057 |
| Unvaccinated |  | 0.30 | 0.20, 0.44 |  | 0.51 | 0.25, 1.00 |  |
| Vaccinated |  | 0.71 | 0.54, 0.92 |  | 1.01 | 0.55, 1.82 |  |
| **Anorexia** | 3,061 |  |  |  |  |  |  |
| No |  | 1.00 | Ref | 0.14 |  |  |  |
| Yes |  | 0.68 | 0.39, 1.10 |  |  |  |  |
| **Dark urine** | 2,952 |  |  |  |  |  |  |
| No |  | 1.00 | Ref | **0.022** |  |  |  |
| Yes |  | 0.55 | 0.32, 0.89 |  |  |  |  |
| **Fatigue** | 3,112 |  |  |  |  |  |  |
| No |  | 1.00 | Ref | **0.016** |  |  |  |
| Yes |  | 1.45 | 1.07, 1.94 |  |  |  |  |
| **Hemorrhagic symptoms** | 2,992 |  |  |  |  |  |  |
| No |  | 1.00 | Ref | **<0.001** |  |  |  |
| Yes |  | 5.18 | 1.83, 12.9 |  |  |  |  |
| **Vomiting** | 3,095 |  |  |  |  |  |  |
| No |  | 1.00 | Ref | **<0.001** | 1.00 | Ref | **0.002** |
| Yes |  | 2.51 | 1.76, 3.52 |  | 2.62 | 1.39, 4.83 |  |

OR = Odds Ratio
CI = Confidence Interval
Statistically significant p-values are in bold text.

Considering the higher YFV positivity rate among vaccinated cases compared to the unvaccinated and the fact that being unvaccinated emerged as a protective factor in our final multivariate logistic regression analysis, we further disaggregated the dataset by vaccination status and analyzed the predictors of YFD by sub-group. The odds of YFD were higher among the unvaccinated and those with unknown vaccination status compared to those with previous YF vaccination (Table 3). Factors associated with YFD among those with unknown vaccination status were the first, third and last quarter of the year compared to the second quarter (OR: 4.27, 95% CI: 2.22–9.26; OR: 9.24, 95% CI: 4.95–19.80, and OR: 24.50, 95% CI: 13.38–51.60, p<0.001); the covid-19 pandemic period (OR: 1.84, 95% CI: 1.51–2.23; p<0.001); the rainy season (OR: 1.39, 95% CI: 1.12–1.71; p=0.003); male sex (OR: 1.45, 95% CI: 1.19–1.78; p<0.001); age being 5 years or older compared to being four years and below (5-9 year: OR: 2.16, 95% CI: 1.33–3.62; 10-14 years: OR: 2.60, 95% CI: 1.57–4.41; 15-29 years: OR: 4.37, 95% CI: 2.88-6.98; 30-44 years: OR: 2.96, 95% CI: 1.86–4.89; 45-59 years: OR: 3.56, 95% CI: 2.07–6.21; p<0.001); residing within the Derived Savannah, the Lowland/Freshwater/Mangrove rainforest, and the Guinea Savannah/Jos Plateau compared to the Sahel/Sudan Savannah (OR: 3.78, 95% CI: 2.80–5.16, OR: 2.01, 95% CI: 1.45–2.83, OR: 4.35, 95% CI: 3.15–6.07 respectively; p<0.001); being unemployed, student, and butcher/hunter/farmer (OR: 1.26, 95% CI: 1.01–1.57; OR: 2.40, 95% CI: 1.69–3.35; and OR: 3.33, 95% CI: 2.15–5.03; p<0.001); working in outdoor settings (OR: 2.21, 95% CI: 1.55–3.17; p<0.001); having traveled within last two weeks preceding onset (OR: 5.53, 95% CI: 3.45–8.65), presenting with fatigue, jaundice, and vomiting (OR: 1.97, 95% CI: 1.38–2.77, p<0.001; OR: 1.50, 95% CI: 1.12–2.02; p=0.007; and OR: 2.37, 95% CI: 1.55–3.56, p<0.001).

**Table 3.** Seasonal, geographical, socio-demographic, clinical factors associated with YFD among reported cases in Nigeria, 2017-2021 disaggregated by vaccination status.

| Characteristic | Vaccinated |  |  |  | Unvaccinated |  |  |  | Unknown status |  |  |  |
| --- | --- | --- | --- | --- | --- | --- | --- | --- | --- | --- | --- | --- |
|  | Total | cOR | 95% CI | p-value | Total | cOR | 95% CI | p-value | Total | cOR | 95% CI | p-value |
| **Annual quarter** | 1,094 |  |  |  | 1,007 |  |  |  | 5,539 |  |  |  |
| First quarter |  | 3.51 | 1.61, 8.80 | **0.017** |  | 1.32 | 0.29, 6.73 | **<0.001** |  | 4.27 | 2.22, 9.26 | **<0.001** |
| Second quarter |  | 1.00 | Ref |  |  | 1.00 | Ref |  |  | 1.00 | Ref |  |
| Third quarter |  | 2.97 | 1.26, 7.79 |  |  | 2.28 | 0.60, 10.9 |  |  | 9.27 | 4.95, 19.8 |  |
| Fourth quarter |  | 2.48 | 0.94, 6.95 |  |  | 13.2 | 4.16, 58.5 |  |  | 24.5 | 13.3, 51.6 |  |
| **Epidemic period** | 1,094 |  |  |  | 1,007 |  |  |  | 5,539 |  |  |  |
| Pre-pandemic |  | 1.00 | Ref | 0.800 |  | 1.00 | Ref | 0.800 |  | 1.00 | Ref | **<0.001** |
| Pandemic |  | 0.94 | 0.56, 1.56 |  |  | 1.10 | 0.50, 2.62 |  |  | 1.84 | 1.51, 2.23 |  |
| **Climatic season** | 1,094 |  |  |  | 1,007 |  |  |  | 5,539 |  |  |  |
| Dry season |  | 1.00 | Ref | 0.900 |  | 1.00 | Ref | 0.800 |  | 1.00 | Ref | **0.003** |
| Rainy season |  | 1.06 | 0.56, 1.88 |  |  | 0.91 | 0.37, 2.05 |  |  | 1.39 | 1.12, 1.71 |  |
| **Geographical region** | 1,094 |  |  |  | 1,007 |  |  |  | 5,539 |  |  |  |
| North |  | 1.00 | Ref | **0.007** |  | 1.00 | Ref | **<0.001** |  | 1.00 | Ref | 0.200 |
| South |  | 2.37 | 1.31, 4.60 |  |  | 6.51 | 2.89, 16.0 |  |  | 1.15 | 0.94, 1.39 |  |
| **Ecological zone** | 1,094 |  |  |  | 1,007 |  |  |  | 5,539 |  |  |  |
| Sahel/Sudan Savannah |  | 1.00 | Ref | **0.002** |  | 1.00 | Ref | **<0.001** |  | 1.00 | Ref | **<0.001** |
| Derived Savannah |  | 2.77 | 0.89, 12.1 |  |  | 48.1 | 9.65, 874 |  |  | 3.78 | 2.80, 5.16 |  |
| Lowland/Freshwater/Mangrove rainforest |  | 6.39 | 2.27, 26.7 |  |  | 19.1 | 3.05, 367 |  |  | 2.01 | 1.45, 2.83 |  |
| Guinea Savannah/Jos Plateau |  | 3.34 | 0.98, 15.3 |  |  | 16.6 | 2.64, 318 |  |  | 4.35 | 3.15, 6.07 |  |
| **Sex** | 1,092 |  |  |  | 1,004 |  |  |  | 5,515 |  |  |  |
| Female |  | 1.00 | Ref | 0.800 |  | 1.00 | Ref | 0.065 |  | 1.00 | Ref | **<0.001** |
| Male |  | 1.08 | 0.65, 1.81 |  |  | 2.40 | 1.00, 6.68 |  |  | 1.45 | 1.19, 1.78 |  |
| **Age group** | 1,012 |  |  |  | 1,000 |  |  |  | 5,309 |  |  |  |
| 0-4 years |  | 1.00 | Ref | **0.031** |  | 1.00 | Ref | **0.045** |  | 1.00 | Ref | **<0.001** |
| 5-9 years |  | 1.15 | 0.42, 3.27 |  |  | 5.03 | 0.80, 97.0 |  |  | 2.16 | 1.33, 3.62 |  |
| 10-14 years |  | 1.37 | 0.46, 4.08 |  |  | 1.28 | 0.05, 32.6 |  |  | 2.60 | 1.57, 4.41 |  |
| 15-29 years |  | 2.58 | 1.13, 6.65 |  |  | 3.88 | 0.70, 72.2 |  |  | 4.37 | 2.88, 6.98 |  |
| 30-44 years |  | 2.88 | 1.12, 7.93 |  |  | 6.16 | 1.04, 117 |  |  | 2.96 | 1.86, 4.89 |  |
| 45-59 years |  | 0.69 | 0.04, 4.03 |  |  | 10.4 | 1.30, 213 |  |  | 3.56 | 2.07, 6.21 |  |
| 60+ years |  | No data |  |  |  | 6.78 | 0.26, 176 |  |  | 1.40 | 0.58, 3.05 |  |
| **Educational status** | 1,094 |  |  |  | 1,007 |  |  |  | 5,539 |  |  |  |
| Educated |  | 1.00 | Ref | 0.600 |  | 1.00 | Ref | 0.140 |  | 1.00 | Ref | **0.002** |
| No education |  | 0.86 | 0.51, 1.52 |  |  | 0.55 | 0.25, 1.24 |  |  | 0.64 | 0.48, 0.86 |  |
| **Occupational group** | 1,094 |  |  |  | 1,007 |  |  |  | 5,539 |  |  |  |
| Home/office worker |  | 1.00 | Ref | **0.019** |  | 1.00 | Ref | **<0.001** |  | 1.00 | Ref | **<0.001** |
| Unemployed |  | 2.09 | 1.13, 3.92 |  |  | 2.13 | 0.59, 7.75 |  |  | 1.26 | 1.01, 1.57 |  |

Table 3 Cont.
| Characteristic | Vaccinated |  |  |  | Unvaccinated |  |  |  | Unknown status |  |  |  |
| --- | --- | --- | --- | --- | --- | --- | --- | --- | --- | --- | --- | --- |
|  | Total | cOR | 95% CI | p-value | Total | cOR | 95% CI | p-value | Total | cOR | 95% CI | p-value |
| Student |  | 1.87 | 0.94, 3.68 |  |  | 3.83 | 1.14, 13.5 |  |  | 2.40 | 1.69, 3.35 |  |
| Butcher/hunter/farmer |  | 1.38 | 0.32, 4.21 |  |  | 6.78 | 2.37, 22.1 |  |  | 3.33 | 2.15, 5.03 |  |
| Artisan/trader |  | 0.50 | 0.03, 2.48 |  |  | No data |  |  |  | 1.81 | 0.90, 3.31 |  |
| Healthcare worker |  | 7.83 | 0.38, 64.3 |  |  | No data |  |  |  | No data |  |  |
| **Occupational setting** | 526 |  |  |  | 604 |  |  |  | 1,084 |  |  |  |
| Indoor |  | 1.00 | Ref | >0.900 |  | 1.00 | Ref | **0.011** |  | 1.00 | Ref | **<0.001** |
| Outdoor |  | 0.98 | 0.42, 2.15 |  |  | 3.43 | 1.36, 9.34 |  |  | 2.21 | 1.55, 3.17 |  |
| **Traveled within last two weeks** | 1,094 |  |  |  | 1,007 |  |  |  | 5,539 |  |  |  |
| No |  | 1.00 | Ref | 0.500 |  | 1.00 | Ref | **0.020** |  | 1.00 | Ref | **<0.001** |
| Yes |  | 2.03 | 0.11, 11.3 |  |  | 6.21 | 0.94, 24.1 |  |  | 5.53 | 3.45, 8.65 |  |
| **Fatigue** | 630 |  |  |  | 647 |  |  |  | 1,835 |  |  |  |
| No |  | 1.00 | Ref | 0.300 |  | 1.00 | Ref | 0.084 |  | 1.00 | Ref | **<0.001** |
| Yes |  | 0.55 | 0.16, 1.44 |  |  | 2.24 | 0.89, 5.73 |  |  | 1.97 | 1.38, 2.77 |  |
| **Jaundice** | 659 |  |  |  | 677 |  |  |  | 1,864 |  |  |  |
| No |  | 1.00 | Ref | 0.130 |  | 1.00 | Ref | 0.130 |  | 1.00 | Ref | **0.007** |
| Yes |  | 1.89 | 0.87, 4.54 |  |  | 0.47 | 0.18, 1.36 |  |  | 1.50 | 1.12, 2.02 |  |
| **Hemorrhagic symptoms** | 618 |  |  |  | 623 |  |  |  | 1,751 |  |  |  |
| No |  | 1.00 | Ref | 0.06 |  | 1.00 | Ref | **<0.001** |  | 1.00 | Ref | 0.120 |
| Yes |  | 9.75 | 0.45, 105 |  |  | 23.6 | 2.96, 151 |  |  | 2.84 | 0.63, 9.39 |  |
| **Vomiting** | 636 |  |  |  | 649 |  |  |  | 1,810 |  |  |  |
| No |  | 1.00 | Ref | **0.044** |  | 1.00 | Ref | **<0.001** |  | 1.00 | Ref | **<0.001** |
| Yes |  | 2.61 | 0.94, 6.27 |  |  | 5.18 | 1.85, 13.6 |  |  | 2.37 | 1.55, 3.56 |  |

cOR = Crude Odds Ratio
CI = Confidence Interval
Vaccinated are cases with previous YF vaccination.
Unvaccinated had no previous YF vaccination.
Unknown status are cases whose YF vaccination history was not known.
Statistically significant p-values are in bold text.

The predictors of YFD among those with previous YF vaccination were the first quarter of the year compared to the second (aOR: 4.04, 95% CI: 1.48–12.95; p=0.010), residing in the southern region (aOR: 14.03, 95% CI: 4.09–88.27; p<0.001). Clinical symptoms such as jaundice (aOR: 2.05, 95% CI: 0.88–5.25; p=0.110) and vomiting (aOR: 2.29, 95% CI: 0.75–6.25; p=0.120) were associated with YFD among the vaccinated but not statistically significant. Predictors among the unvaccinated were male sex (aOR: 6.35, 95% CI: 1.66–32.16; p=0.012), residing in the southern region (aOR: 76.64, 95% CI: 13.57–1465.89; p<0.001), working in outdoor settings (aOR: 3.11, 95% CI: 0.91–12.03; p=0.079), presenting with fatigue (aOR: 3.87, 95% CI: 1.10–15.08; p=0.039), hemorrhagic symptoms (aOR: 368.00, 95% CI: 14.29–14759.72; p<0.001), and vomiting (aOR: 6.53, 95% CI: 1.57–27.23; p= 0.009). Predictors of YFD among cases with unknown vaccination status were the third and fourth quarter of the year (aOR: 4.44, 95% CI: 1.92–12.11; aOR: 3.38, 95% CI: 1.08–10.90; p=0.001); age being 15 years or older (aOR: 1.87, 95% CI: 1.17–3.10, p=0.012);and residing within the Derived Savannah and the Guinea Savannah/Jos Plateau (aOR: 7.09, 95% CI: 3.35–17.45 and aOR: 5.24, 95% CI: 2.41–13.14 respectively; p<0.001) Fig 6.

**Fig 6.**
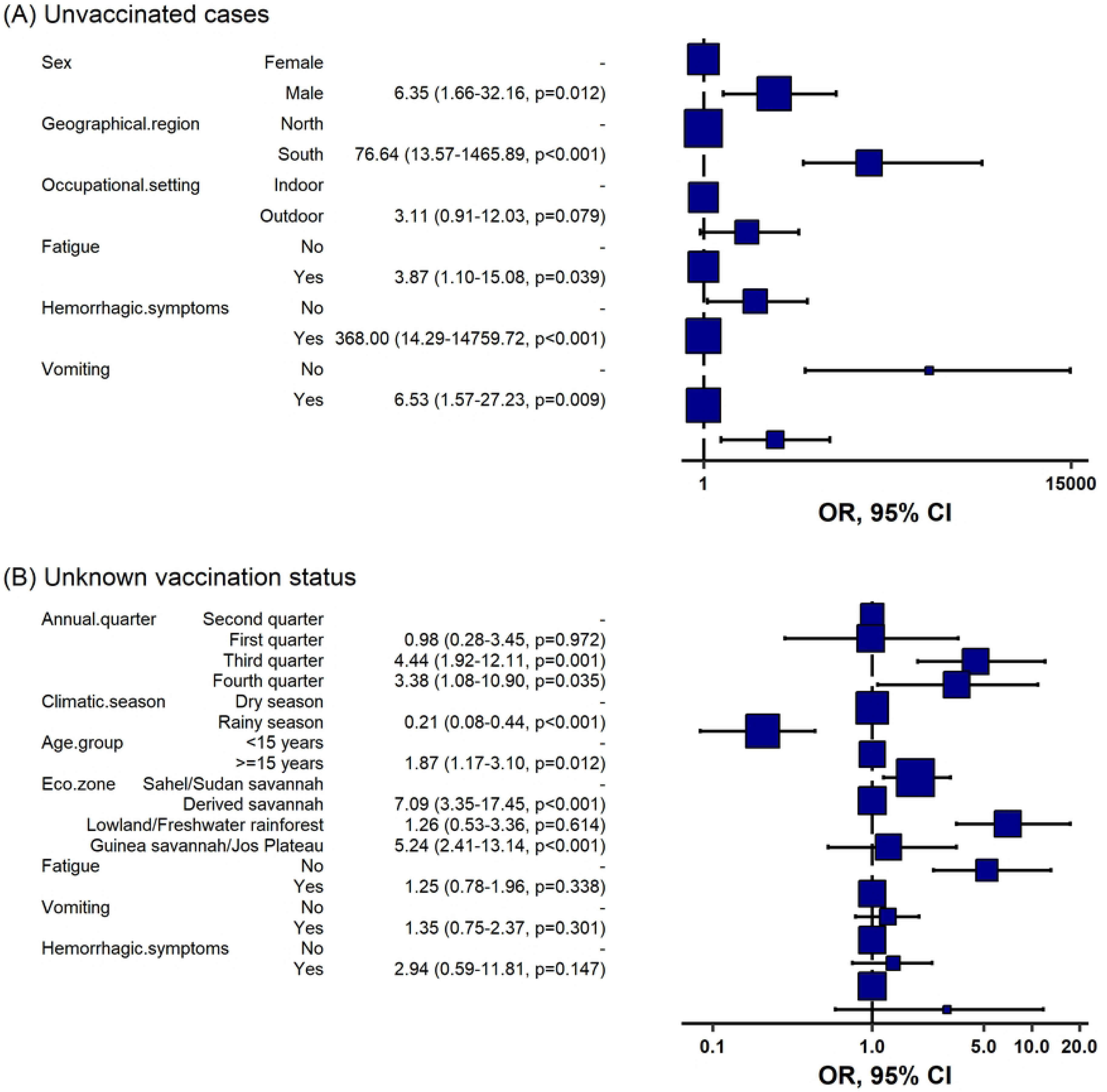
Seasonal, geographical, socio-demographic, clinical predictors of YFD among reported cases in Nigeria, 2017-2021 disaggregated by vaccination status. (A) Unvaccinated (B) Unknown vaccination status. The first column shows the explanatory variables, second column show the levels and third column is the adjusted odds ratios with corresponding confidence intervals.

### Factors associated with being unvaccinated (barriers to YF vaccination)

Information on vaccination status was available for 3983 out 13014 reported cases, of which 1735 (13.3%) were vaccinated, 2248 (17.3) were unvaccinated, and 9031 (69.4%) were of unknown status (Table 1). The most unvaccinated age group were those 15-29 years, which also accounted for 30.0% of the total population, 32.0 % of unvaccinated cases, and 27.0% of cases of unknown vaccination status (Fig 4 and S1 Table). A comparison of the characteristics of the cases by vaccination status is shown in S1 Table. Independent predictors of being unvaccinated for YF, irrespective of laboratory test status, were the rainy season compared to the dry season (aOR: 1.29, 95% CI: 1.11-1.66, p=0.003); being 15years or older compared to under-five (15-29 years: aOR: 2.06, 95% CI: 1.51-2.83; 30-44 years: aOR: 2.11, 95% CI: 1.45-3.07; 45-59 years: aOR: 2.72, 95% CI: 1.63-4.58; 60+ years: aOR: 6.55, 95% CI: 2.76-17.50; p<0.001); residing in the northern region compared to the southern region (aOR: 3.71, 95% CI: 3.01-4.58, p<0.001); and occupation being either butcher, farmer or hunter compared to being home-based/officer worker (aOR: 2.30, 95% CI: 1.52-3.50, p<0.001). Working in outdoor setting was protective against being unvaccinated (aOR: 0.46, 95% CI: 0.31-0.69, p<0.001) (Table 4).

**Table 4.** Seasonal, geographical, and socio-demographic factors associated with non-vaccination among reported YF cases in Nigeria, 2017-2021.

|  | Univariate |  |  |  | Multivariable |  |  |
| --- | --- | --- | --- | --- | --- | --- | --- |
| Characteristic | Total | Crude OR | 95% CI | p-value | Adjusted OR | 95% CI | p-value |
| **Epidemic period** | 3,983 |  |  |  |  |  |  |
| Pre-pandemic |  | 1.00 | Ref | 0.400 |  |  |  |
| Pandemic |  | 1.06 | 0.93, 1.20 |  |  |  |  |
| **Climatic season** | 3,983 |  |  |  |  |  |  |
| Dry season |  | 1.00 | Ref | 0.063 | 1.00 | Ref | **0.013** |
| Rainy season |  | 1.15 | 0.99, 1.33 |  | 1.29 | 1.05, 1.57 |  |
| **Sex** | 3,971 |  |  |  |  |  |  |
| Female |  | 1.00 | Ref | 0.200 |  |  |  |
| Male |  | 1.09 | 0.96, 1.24 |  |  |  |  |
| **Age group** | 3,862 |  |  |  |  |  |  |
| 0-4 years |  | 1.00 | Ref | **<0.001** | 1.00 | Ref | **<0.001** |
| 5-9 years |  | 0.85 | 0.69, 1.05 |  | 0.92 | 0.66, 1.28 |  |
| 10-14 years |  | 1.08 | 0.86, 1.37 |  | 1.25 | 0.86, 1.82 |  |
| 15-29 years |  | 1.63 | 1.34, 1.97 |  | 2.06 | 1.51, 2.83 |  |
| 30-44 years |  | 1.59 | 1.27, 2.00 |  | 2.11 | 1.45, 3.07 |  |
| 45-59 year |  | 1.58 | 1.13, 2.22 |  | 2.72 | 1.63, 4.58 |  |
| 65+ years |  | 2.28 | 1.38, 3.89 |  | 6.55 | 2.76, 17.5 |  |
| **Geographical region** | 3,983 |  |  |  |  |  |  |
| South |  | 1.00 | Ref | **<0.001** | 1.00 | Ref | **<0.001** |
| North |  | 4.66 | 4.07, 5.35 |  | 3.71 | 3.01, 4.58 |  |
| **Educational status** | 3,983 |  |  |  |  |  |  |
| Educated |  | 1.00 | Ref | <0.001 | 1.00 | Ref | 0.700 |
| No education |  | 1.39 | 1.20, 1.59 |  | 1.04 | 0.82, 1.33 |  |
| **Occupational status** | 3,983 |  |  |  |  |  |  |
| Home/office worker |  | 1.00 | Ref | **<0.001** | 1.00 | Ref | **<0.001** |
| Unemployed |  | 1.00 | 0.85, 1.17 |  | - | - |  |
| Student |  | 0.54 | 0.45, 0.64 |  | 0.62 | 0.47, 0.83 |  |
| Butcher/hunter/farmer |  | 1.91 | 1.51, 2.42 |  | 2.30 | 1.52, 3.50 |  |
| Artisan/trader |  | 0.80 | 0.58, 1.11 |  | 1.45 | 0.90, 2.37 |  |
| Healthcare worker |  | 2.12 | 0.98, 5.09 |  | 1.42 | 0.63, 3.54 |  |
| **Occupational setting** | 2,119 |  |  |  |  |  |  |
| Indoor |  | 1.00 | Ref | **0.003** | 1.00 | Ref | **<0.001** |
| Outdoor |  | 1.32 | 1.10, 1.59 |  | 0.42 | 0.28, 0.62 |  |

OR = Odds Ratio
CI = Confidence Interval
Statistically significant p-values are in bold text.

To further investigate the effect of the COVID-19 pandemic on YF vaccination, we disaggregated the analysis by laboratory test status. The COVID-19 pandemic period was associated with being unvaccinated among YFV positive (OR: 2.43, 95% CI: 0.96-6.48, p=0.066) and negative cases (OR: 2.06, 95% CI: 1.73-2.47, p<0.001) compared to the pre-pandemic period, just as working in outdoor settings in comparison to working indoors (OR: 3.81, 95% CI: 1.16-13.6, p=0.032) among YFV positive cases (Table 5).

**Table 5.** Factors associated with non-vaccination among reported Yellow fever cases in Nigeria, 2017-2021 disaggregated by laboratory test status.

| Characteristic | Positive cases |  |  |  | Negative cases |  |  |  | Unconfirmed cases |  |  |  |
| --- | --- | --- | --- | --- | --- | --- | --- | --- | --- | --- | --- | --- |
|  | N | cOR | 95% CI | p-value | N | cOR | 95% CI<sup>1</sup> | p-value | N | cOR | 95% CI | p-value |
| **Epidemic period** | 90 |  |  |  | 2,011 |  |  |  | 442 |  |  |  |
| Pre-pandemic |  | 1.00 | Ref | 0.066 |  | 1.00 | Ref | **<0.001** |  | 1.00 | Ref | 0.120 |
| Pandemic |  | 2.43 | 0.96, 6.48 |  |  | 2.06 | 1.73, 2.47 |  |  | 0.34 | 0.22, 0.54 |  |
| **Climatic season** | 90 |  |  |  | 2,011 |  |  |  | 442 |  |  |  |
| Dry season |  | 1.00 | Ref |  |  | 1.00 | Ref |  |  | 1.00 | Ref | 0.060 |
| Rainy season |  | 1.45 | 0.51, 3.96 | 0.500 |  | 1.69 | 1.39, 2.06 | **<0.001** |  | 0.59 | 0.34, 1.03 |  |
| **Geographical region** | 90 |  |  |  | 2,011 |  |  |  | 442 |  |  |  |
| South |  | 1.00 | Ref | 0.300 |  | 1.00 | Ref | **<0.001** |  | 1.00 | Ref | **<0.001** |
| North |  | 1.74 | 0.60, 4.86 |  |  | 4.79 | 3.96, 5.80 |  |  | 6.71 | 4.23, 10.8 |  |
| **Sex** | 88 |  |  |  | 2,008 |  |  |  | 439 |  |  |  |
| Female |  | 1.00 | Ref | 0.110 |  | 1.00 | Ref | 0.600 |  | 1.00 | Ref | 0.100 |
| Male |  | 2.33 | 0.85, 7.14 |  |  | 1.05 | 0.88, 1.25 |  |  | 1.43 | 0.93, 2.19 |  |
| **Age group** | 83 |  |  |  | 1,929 |  |  |  | 1,850 |  |  |  |
| 0-4 years |  | 1.00 | Ref | 0.053 |  | 1.00 | Ref | **<0.001** |  | 1.00 | Ref | **<0.001** |
| 5-9 years |  | 3.89 | 0.47, 83.8 |  |  | 0.89 | 0.66, 1.19 |  |  | 0.82 | 0.60, 1.11 |  |
| 10-14 years |  | 1.00 | 0.03, 28.8 |  |  | 1.07 | 0.78, 1.48 |  |  | 1.21 | 0.85, 1.73 |  |
| 15-29 years |  | 2.55 | 0.36, 51.5 |  |  | 1.70 | 1.30, 2.22 |  |  | 1.67 | 1.26, 2.22 |  |
| 30-44 years |  | 3.50 | 0.46, 73.3 |  |  | 1.64 | 1.19, 2.26 |  |  | 1.67 | 1.18, 2.36 |  |
| 45-59 years |  | 21.0 | 1.33, 913 |  |  | 1.39 | 0.87, 2.25 |  |  | 1.68 | 1.03, 2.81 |  |
| 60+ years |  |  | No data |  |  | 2.03 | 1.02, 4.17 |  |  | 2.48 | 1.16, 5.95 |  |
| **Educational status** | 90 |  |  |  | 2,011 |  |  |  | 1,882 |  |  |  |
| Educated |  | 1.00 | Ref | 0.300 |  | 1.00 | Ref | 0.800 |  | 1.00 | Ref | **<0.001** |
| No education |  | 0.62 | 0.24, 1.61 |  |  | 0.98 | 0.80, 1.19 |  |  | 2.10 | 1.70, 2.60 |  |
| **Occupational group** | 90 |  |  |  | 2,011 |  |  |  | 442 |  |  |  |
| Home-based/Office worker |  | 1.00 | Ref | **0.002** |  | 1.00 | Ref | **<0.001** |  | 1.00 | Ref | **<0.001** |
| Unemployed |  | 0.87 | 0.21, 3.55 |  |  | 0.85 | 0.68, 1.07 |  |  | 2.02 | 1.11, 3.83 |  |
| Student |  | 1.50 | 0.38, 6.09 |  |  | 0.73 | 0.57, 0.94 |  |  | 0.21 | 0.11, 0.41 |  |
| Butcher/farmer/hunter |  | 13.3 | 2.92, 79.5 |  |  | 2.72 | 1.93, 3.88 |  |  | 0.86 | 0.45, 1.73 |  |
| Artisan/Trader |  | Missing |  |  |  | 0.90 | 0.58, 1.40 |  |  | 0.38 | 0.11, 1.37 |  |
| Healthcare worker |  | Missing |  |  |  | 6.70 | 2.27, 28.7 |  |  | 0.32 | 0.01, 8.10 |  |
| **Occupational setting** | 48 |  |  |  | 1,082 |  |  |  | 225 |  |  |  |
| Indoor |  | 1.00 | Ref | **0.032** |  | 1.00 | Ref | 0.500 |  | 1.00 | Ref | 0.150 |
| Outdoor |  | 3.81 | 1.16, 13.6 |  |  | 1.09 | 0.85, 1.41 |  |  | 1.52 | 0.87, 2.70 |  |

cOR = Crude Odds Ratio
CI = Confidence Interval
Positive cases are YFV laboratory confirmed cases of YF.
Negative cases are suspected cases that tested negative for YFV.
Unconfirmed cases are suspected cases that had no laboratory confirmation.
Statistically significant p-values are in bold text.

More independent predictors were seen among YFV negative cases, which include the COVID- 19 pandemic period (aOR: 1.40, 95% CI: 1.14-1.72, p=0.002), age group being 15-29, 30-44, 60+ years (aOR: 1.81, 95% CI: 1.28-2.57; aOR: 1.79, 95% CI: 1.19-2.69; and aOR: 2.25, 95% CI: 1.03-5.02; p=0.001), residing in the northern region (aOR: 4.39, 95% CI: 3.58-5.40, p<0.001); and occupation being butcher/farmer/hunter, and health worker (aOR: 1.63, 95% CI: 1.08-2.47; aOR: 4.03, 95% CI: 1.33-17.47; p=0.021) (**Fig 7**).

**Fig 7.**
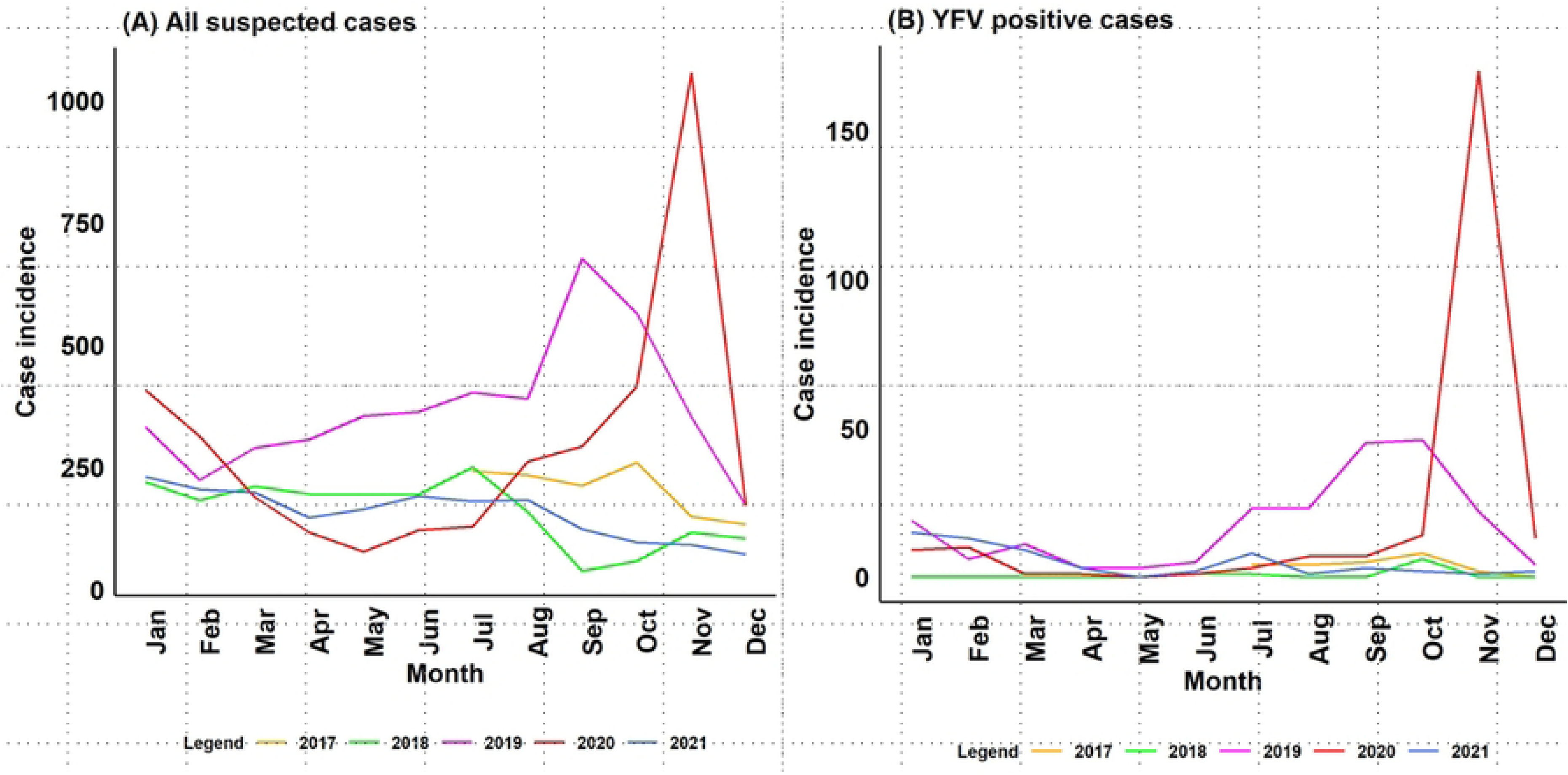
Seasonal, geographical, and socio-demographic predictors non-vaccination among reported cases in Nigeria, 2017-2021 disaggregated by laboratory test status. (A) Negative (B) Unconfirmed cases. The first column shows the explanatory variables, second column show the levels and third column is the adjusted odds ratios with corresponding confidence intervals.

Among YFV positive cases, independent predictors of being unvaccinated were COVID-19 pandemic period (aOR: 3.08, 95% CI: 0.86-12.55, p=0.09) and working in outdoor settings (aOR: 3.89, 95% CI: 1.13-14.64, p=0.03).

## Discussion

Recommended control strategies for YF under the WHO EYE strategy include effective and timely surveillance, strengthening laboratory services, adequate routine immunization coverage and a comprehensive response to outbreaks of YF [11,17,29,30]. In Nigeria, YF surveillance is a passive, health facility-based, case-based, laboratory-based, surveillance system [18]. Most suspected cases reported during the study period were not confirmed by laboratory test, which may be because cases did not present early enough at health facilities to enable prompt detection of YFV by RT-PCR within the ten-day window period for detection of the viral genome in blood samples. Logistical challenges associated with sample transportation from Nigeria to the Regional Reference Laboratory in Dakar, Senegal for confirmation of presumptive positive and inconclusive RT-PCR negative samples by PRNT may be another possible reason [18]. Late presentation of cases at health facility may have also contributed to the large proportion of suspected cases that tested negative for YFV despite meeting the case definition. These challenges may be surmounted by investment in community-based surveillance to enable prompt case detection and building in-country capacity for full complement of serological testing. In Côte d’Ivoire, an eight-fold increase in suspected YF case reporting was documented following implementation of community-based surveillance [31].

The odds of YFD increased significantly from the 2017 epidemic season through the 2020 season, which was followed by an abrupt decrease in the 2021 season. The 2020 epidemic season coincided with the commencement of the COVID-19 pandemic. Thus, the decrease in case reporting observed during the pandemic period may be a pointer to the disruptive effect of the COVID-19 pandemic on YF surveillance activities in Nigeria [32]. The pandemic resulted in the conversion of several public hospitals to COVID-19 treatment centers in Nigeria and elicited patients’ apathy towards hospital attendance [33]. We have shown that cases were about twice as likely to be unvaccinated for YF during the pandemic period. Ayobami et al (2023) had reported that the COVID-19 pandemic was associated with reduction in care seeking for children and clinic attendance for childhood immunization [33]. Similarly, Ribeiro da Silva et al (2022) reported a reduction in the number of YF vaccine doses administered in Brazil during the pandemic period compared to the pre-pandemic era [34].

Our study has shown that YF outbreaks in Nigeria followed a distinct seasonal pattern in which case incidence followed the rainy season progression. Previous studies in Uganda and Brazil have reported a rainy season driven YF outbreak pattern [35,36]. The YF mass vaccination campaigns implemented in Nigeria between 2017 and 2019 were carried out in September of each year [5]. We showed that cases were nearly twice more likely to be unvaccinated during the rainy season. The rainy season is the farming season in Nigeria, and farmers tend to spend most of their daytime in their farms during this time of year. Perhaps, commencing reactive vaccination early in the year (January - April) may increase the chances of getting more persons vaccinated, especially in areas where people are predominantly farmers or get involved in other forest-going occupation.

The most-at-risk population in this study were younger people, especially males. A similar study in Brazil reported males with a median age of 26 (range: 1-93) years, with the most affected age group being 20-29 years [23]. Ribeiro et al. also found males with a mean age of 29 years [37]. We found that cases in the age group 15-29 years had over two-fold odds of being unvaccinated and over four-fold odds of having YFD. Higher odds of YFD in males compared to females seen in this study has been attributed to occupation. A study in Uganda found that cases were predominantly males involved in rural agricultural activities in forested, swampy areas - an observation that has also been reported in Ethiopia [4,35].

The finding that cases without any travel history featured prominently in the epidemic suggests the presence of suitable conditions for YFV transmission locally [6]. However, persons with travel history had over five-fold odds of YFD compared to those that did not. Studies in Uganda [35] and Brazil [23] have incriminated traveling to YF endemic areas as risk factor for infection. This observation may not be unconnected with ecological characteristics. We found that the north-west geo-political zone, which straddles parts of the Guinea Savannah, Jos Plateau, Sahel Savannah, and the Sudan Savannah, reported the highest proportion of suspected cases but YFV positivity was highest in the south-east, which occupies parts of the Derived Savannah and the Lowland rainforest. Nomhwange et al (2021) had previously identified four epidemiological blocks of YFV spread in Nigeria, delineated by the geo-political zones: block 1 around the north-central; block 2 in the south-south; block 3 in the south-east; and block 4 in the north-east [5]. The outbreak in the north-central block has been reported to spread into the north-west while the south-south block spreads into the south-west. Our findings, however, suggest that YFV transmission in these epidemiological blocks seemed to be related to the characteristics of the ecological zones that occupy the geo-political zones. We demonstrated for the first time, to our knowledge, the important contribution of the Derived Savannah, the Lowland/Freshwater/Mangrove rainforest, and the Guinea Savannah/Jos Plateau to YF epidemiology in Nigeria. Of concern is the double jeopardy of the higher odds YFD among unvaccinated and vaccinated persons in the Lowland/Freshwater/Mangrove rainforest ecological zone, which mainly occupy the south-south and south-west geo-political zones. We further showed that the odds of YFD in the Derived Savannah and the Guinea Savannah/Jos Plateau could be mitigated by adequate vaccination as these ecological zones largely lie in the northern region where cases had about five-fold odds of being unvaccinated compared to those in the southern region. A combination of human, reservoir, and vector surveillance studies, in addition to the effects of human encroachment may provide further insight as to whether the varying degree of risk of YFD seen across the ecological zones in this study is driven by specific characteristics. In Brazil, where the YFV has demonstrated its ability to expand its geographic spread since 2016, human activities have been implicated as contributing factor [36,38].

Clinically, the viraemia phase of YF is characterized by fever, headache, chills, backache, fatigue, anorexia, nausea, and vomiting, whereas the toxaemic phase is marked by high fever, worsening headache, chills, muscle pain and signs of multiple organs and systems involvement. Hepatic coagulopathy leads to severe hemorrhagic manifestations [6]. These symptoms were manifested by the cases reported in this study, just as had also been reported in Uganda [35] and Brazil [2,39]. YF is distinguished from other hemorrhagic fevers caused by viruses by the characteristic liver damage and presentation of jaundice [6]. Presenting fatigue, hemorrhagic symptoms and vomiting were significantly associated with YFD among the unvaccinated in our study. These factors may be important in strengthening YF case definition for the purpose of surveillance as well enhancing index of suspicion by clinicians and surveillance officers.

Vaccination is widely recognized as the most potent weapon to combat YF outbreaks. Following the resurgence of YF outbreak in 2017 in Nigeria, efforts were made to revamp up YF vaccination coverage. A total 45,648,243 persons of age less than 45 years were vaccinated across Nigeria through nine targeted reactive mass vaccination campaigns and three phased mass vaccination activities. [5]. The coverage achieved by these campaigns has not proved to adequately protect the population against YFV spread. In developing a strategy to enhance vaccine uptake in Nigeria, it is imperative that policy makers take into cognizance the potential barriers to YF vaccination. The independent predictors of being unvaccinated were the COVID-19 pandemic period, rainy season, being 15 years or older, residing in northern Nigeria, being a butcher, farmer, or hunter and being a healthcare worker. To meet the 2026 target of the WHO EYE strategy, a review of the current policy on vaccination may be warranted to achieve sufficient coverage that could halt YFV transmission in Nigeria. A sizeable proportion of vaccinated cases were found to be positive for YFV and hospitalized, pointing to evidence of possible vaccine failure. A similar finding was reported by Tuboi et al (2007) in Brazil [23]. It may be difficult to pinpoint the reason for this. It is, however, known that the 17D YF vaccine is a live attenuated vaccine, and it is possible that its efficacy may be affected by factors related to proper handling, transport, storage, and usage, which could be challenging in resource-poor settings [23,40]. The involvement of vaccinated cases in the epidemic coincided with the period of implementation of reactive YF vaccination campaigns in Nigeria. The highest incidence was seen during the 2019 epidemic season, largely among cases 15-29 year, working in indoor setting and residing largely in the Derived Savannah, Lowland rainforest, and the Sudan Savannah. This brings to the fore the crucial need for invest investments research tools that would enable understanding of the molecular epidemiology of the YFV in Nigeria.

### Limitation

This study represents the first attempt in Nigeria to use multi-season national surveillance data to characterize the seasonal, geographical, socio-demographic, and clinical characteristics associated with YFD and to identify barriers to vaccination. The dataset was generated from different health facilities across Nigeria, making it quite representative. However, just like every other surveillance system, the national YF surveillance in Nigeria is not without limitations. First, it is a passive, health facility-based surveillance system, which relies on the judgement of clinicians to identify cases. Although, under outbreak conditions, an active and community-based component of the surveillance system is activated. Considering the broad range of clinical presentation and symptoms associated with YF and the passive nature of the surveillance system, less severe cases may have been missed. However, clinicians and focal persons were trained regularly on using a WHO standard case definition to identify YF cases. Second, the national line list does not capture information on co-morbidities and pre-existing illnesses such as liver disease or previous toxic exposures such as alcohol consumption that may be confounding factors in YF disease. Third, there were issues related to data quality, including missing data, however, the development and deployment of the SORMAS platform, has strengthened the surveillance of diseases captured under the IDSR system, including YF in Nigeria, providing real time data entry and analysis. In any case, secondary data is useful for planning preventive measures, healthcare assessment, and research[41]. Findings from our study has the potential to be generalized as the dataset used had national representation, covering the six geo-political zones in Nigeria. However, it is important to note that the dataset used for the study was largely hospital-based. This implies that cases who felt they were not ill enough to access care in a health facility setting were completely missed by this study.

## Conclusion

In conclusion, following its resurgence in 2017, YF outbreaks in Nigeria occurred annually with increasing odds of incidence until the 2020 epidemic season. Despite the epidemic following the rainy season pattern with the peak incidence seen between September and November annually, the odd of YFD was higher during the dry season. A combination of factors has proven to be important drivers of YF outbreaks in Nigeria. Although vaccination has proven to reduce the odds of YFD, the higher odds of YFD among previously vaccinated persons, especially those residing in the southern region suggests that inadequate vaccination alone may not explain the recurrent YF incidence in Nigeria, warranting further research using genomic surveillance tools. To improve YF vaccine uptake, current policy on vaccination campaigns and routine immunization may need to be reviewed to address the barriers to YF vaccination identified in this study. The COVID-19 pandemic, the rainy season, being 15 years or older, residing in the northern region, and certain occupation have been shown to be impediment to YF vaccination. Our findings are critical for planning public health interventions and to guide research strategies that would enable Nigeria to meet the 2026 target of the WHO EYE strategy.

## Supporting information

**S1 Fig: Map of Nigeria.** (a) geo-political zones (b) ecological zones

**S1 Table. Comparison of seasonal, geographical, and socio-demographic characteristics among reported Yellow fever cases in Nigeria, 2017-2021 by vaccination status.**

## Acknowledgement

We sincerely appreciate the Program for Nurturing Global Leaders in Tropical and Emerging Communicable Diseases, Graduate School of Biomedical Sciences, Nagasaki University, the Management of the Nigeria Centre for Disease Control, and Japan International Cooperation Agency.

## Author contributions

**Conceptualization:** Stephen E. Akar, Kaneko Satoshi

**Data curation:** Stephen E. Akar, Akanimo Iniobong, William Nwachukwu, Chinwe Ochu, Olajumoke Babatunde, Ifedayo Adetifa, Chikwe Ihekweazu

**Data analysis:** Stephen E. Akar, Kaneko Satoshi

**Methodology:** Stephen E. Akar, Mami Hitachi, Kentaro Kato, Kaneko Satoshi

**Supervision:** Satoshi Kaneko, Yuki Takamatsu, Kenji Hirayama

**Validation:** Satoshi Kaneko, Kentaro Kato, Mami Hitachi, Stephen E. Akar

**Writing original (original draft):** Stephen E. Akar, Satoshi Kaneko

Writing (review and editing): Stephen E. Akar, William Nwachukwu, Akanimo Iniobong, Oyeladun Okunromade, Chinwe Ochu, Olajumoke Babatunde, Ifedayo Adetifa, Chikwe Ihekweazu, Mami Hitachi, Kentaro Kato, Yuki Takamatsu, Kenji Hirayama, Satoshi Kaneko

## Data Availability Statement

The dataset used in our study is a third-party data archived with the NCDC, which is the national public health institute in Nigeria. Access is granted by the NCDC on request for specific dataset. Such request could be directed to.

